# Impact of Omicron Wave and Associated Infection Prevention and Control Measures in Shanghai on Health Management and Psychosocial Well-Being of Patients with Chronic Conditions

**DOI:** 10.1101/2022.10.03.22280646

**Authors:** Zhimin Xu, Gabriela Lima de Melo Ghisi, Xia Liu, Lixian Cui, Sherry L Grace

## Abstract

**Background:** COVID-19 and associated controls may be particularly problematic in the context of chronic conditions. This study investigated health management, well-being, and pandemic-related perspectives in these patients in the context of stringent measures, and associated correlates.

**Methods:** A self-report survey was administered via Wenjuanxing in Simplified Chinese between March-June 2022 during the Omicron wave lockdown in Shanghai, China. Items from the Somatic Symptom Scale (SSS) and Symptom Checklist-90 (SCL-90) were administered, as well as pandemic-related items created by a working group of the Chinese Preventive Medical Association. Chronic disease patients in this cross-sectional study were recruited through an associated community family physician group.

**Results:** Overall, 1775 patients, mostly married females with hypertension, participated. Mean SSS scores were 36.1±10.5/80, with 41.5% scoring in the elevated range (i.e., above 36). In an adjusted model, female, diagnosis of coronary artery disease and arrhythmia, perceived impact of pandemic on life, duration can tolerate control measures, perception of future & control measures, impact of pandemic on health condition and change to exercise routine due to pandemic were significantly associated with greater distress. Approximately one-quarter (24.5%) perceived the pandemic had a permanent impact on their life, and 44.1% perceived at least a minor impact on their health. One-third (33.5%) discontinued exercise due to the pandemic. While 47.6% stocked up on their medications before the lockdown, their remaining supply was mostly only enough for a couple of weeks and 17.5% of participants discontinued use. Chief among their fears were inability to access healthcare (83.2%), and what they stated they most needed to manage their condition was medication access (65.6%).

**Conclusions:** Since 2020 when we assessed a similar cohort, distress and perceived impact of the pandemic has worsened. Greater access to cardiac rehabilitation in China could address these issues.

## Background

The Coronavirus Disease (COVID-19) pandemic, with its many unpredictable waves, has resulted in major negative impacts for economies, health systems and citizens worldwide.^1,2^ While the impact of COVID-19 on the health of all people has been of major concern, it has been a particular concern for those at higher risk of severe outcomes. This includes notably patients with chronic conditions such as cardiovascular diseases (CVD). For instance, hospitalization rates and mortality rates are much higher in this population; in the United States alone chronic diseases account for 75% of aggregate healthcare spending and are responsible for 7 out of 10 deaths;^3^ in China, the deaths caused by chronic diseases account for 87% of total deaths.^4^

Measures to control infection spread such as physical distancing often necessitating closure of essential businesses,^5^ as well as diversion of healthcare resources for COVID-19,^6^ have also had a negative impact. In the general population, this has resulted in reduced access to care^7^ as well as increases in mental health conditions.^8^ Despite their greater risk, there is only limited study of the impacts of the pandemic and associated control measures on the psychosocial well-being and self-management of patients with chronic conditions; we identified some abstracts only to date.^9,10^

Government policy varies worldwide in terms of implementation of infection prevention and control measures. Based on the COVID-19 stringency index, such strategies are among the most stringent in China.^11^ For example, in one of the world’s most populous cities of Shanghai, a strict “closed-loop” control system has been enforced. This involves home isolation except for medical reasons, with extra-household interpersonal contact and outdoor time forbidden, and transactions quickly moving online. This caused residents to become anxious and unsettled.^12^ Therefore, the objective of this study was to assess the impacts of strict control measures during the most recent Omicron wave in Shanghai on chronic disease patients in terms of: (1) psychosocial well-being, (2) health management, and (3) perceptions related to the pandemic and associated control measures. The associations of their health / well-being with perceived impact of the pandemic on their health were also tested, as were the association of pandemic-related attitudes with psychological well-being.

## Methods

### Design and Procedure

Study approval was secured from the Ethics Committee of Xinhua Hospital, affiliated with Shanghai Jiaotong University School of Medicine (XHEC-C-2022-042). The anonymous online survey in Simplified Chinese was created by the Cardiac Rehabilitation (CR) Group, Health Risk Assessment and Control of the Chinese Preventive Medical Association. Data collection for this cross-sectional study was undertaken between March-June 2022. It was distributed via Wenjuanxing by family doctors who belonged to the Community Physician Group of the Psychosomatic Medicine Special Committee of the Shanghai Association of Integrated Chinese and Western Medicine; this group cooperates closely with the Chinese Preventive Medical Association.

### Setting and Participants

During the Omicron wave lockdown in Shanghai, residents including patients with chronic conditions were to remain indoors and could not go outside to exercise or purchase groceries at the supermarket. They could receive care via telehealth or consultation in internet hospitals. Medication was available through online pharmacies with in-home delivery. This could be challenging for seniors who may have low digital literacy or who do not have access to a smartphone.

To receive in-person care, health codes had to be scanned. Daily nucleic acid tests were mandatory for in-patients and the one allowed family member who could accompany a patient. Participant inclusion criteria were the following: outpatients with cardiovascular and cerebrovascular diseases, diabetes, kidney or another chronic condition (e.g., cancer). Participant exclusion criteria were: (1) outpatients with severe cognitive impairments or any conditions that prevent them from being able to understand and agree to participate in the research; (2) outpatients unwilling to cooperate with the research protocol; (3) outpatients with any missing data within questionnaire items.

### Measures

All items were self-reported. Five items from the validated Symptom Checklist-90 (SCL-90) were administered to assess depressive symptoms.^13,14^ There is a 5-point Likert-type response option for each, with a maximal score of 25; higher scores indicate more severe symptoms.

The Chinese version of the Somatic Self-rating Scale (SSS) was also administered.^15^ It comprises of 20 items, each assessed on a 4-point scale. Items assess physical (i.e., half the items query each body system), depressive, and anxiety disorders and a final 2 items assess sleep and cognitive issues. Thus, the SSS is more than a measure of somatic symptoms but assesses psychosocial well-being more broadly. The total score can range from 20 to 80; scores above 36 were considered elevated.

In addition, non-psychometrically validated items related to pandemic perceptions were generated by the CR group. Participants were asked to respond these items based on their experience in the prior two weeks.

### Statistical Analysis

IBM SPSS statistics version 25.0 was used for all statistical analysis, with p < 0.05 considered statistically significant. Descriptive analysis was first performed. Perceptions of impact of the pandemic (at least minor vs none) on their chronic condition and psychosocial well-being were tested using chi-square and t-tests as applicable. The association of pandemic-related perceptions with SSS scores was assessed using t-test and chi-square, as applicable.

Finally, associations between participant sociodemographic and clinical characteristics with SSS scores were tested using logistic regression analysis as applicable. Then, an adjusted model was computed for the association of the SSS with those sociodemographic, clinical, and pandemic-related perceptions that were significant at p < 0.05.

## Results

Of 1775 responding patients, most were educated, working, married females living with family, who had healthcare insurance (Table 1). As shown in Table 2, most participants had hypertension, over one-fifth had diabetes, and just under that coronary artery disease. Other conditions included cancer, as well as endocrine and immune diseases, and psychiatric conditions.

**Table 1:** Participant Sociodemographic Characteristics (N=1775)

| Characteristic | n (%) / mean ± SD |
| --- | --- |
| Sex (female) | 1061 (59.8) |
| Age | 57.69±13.80 |
| Marital Status |  |
| *Married* | 1489 (83.9) |
| *Widowed* | 111 (6.3) |
| *Never married* | 107 (6.0) |
| *Separated / divorced* | 68 (3.8) |
| Living Situation |  |
| *With children and/or other extended family members* | 786 (44.3) |
| *With partner only* | 768 (43.3) |
| *Alone* | 221 (12.5) |
| Highest Educational Attainment |  |
| *Junior college or senior high school* | 818 (46.1) |
| *Bachelor and above* | 723 (40.7) |
| *Junior high school and below* | 234 (13.2) |
| Income Status |  |
| *Has work income* | 1501 (84.6) |
| *No fixed income (e.g., rely on children, state subsidies)* | 138 (7.8) |
| *Has additional source of income on top of salary (e.g., online)* | 136 (7.7) |
| Current or Previous Occupation |  |
| *City worker* | 557 (31.4) |
| *Civil servants* | 440 (24.8) |
| *Management (private or public)* | 289 (16.3) |
| *Health care provider* | 181 (10.2) |
| *Teacher* | 129 (7.3) |
| *Government* | 59 (3.3) |
| *Own business* | 44 (2.5) |
| *Farmer* | 34 (1.9) |
| *Research* | 34 (1.9) |
| *Other (e.g., Arts)* | 8 (0.5) |
| Healthcare Insurance |  |
| *Employee* | 960 (54.1) |
| *Town residents* | 664 (37.4) |
| *Off-site* | 77 (4.3) |
| *Self-pay* | 74 (4.2) |
| SD, standard deviation |  |

**Table 2:** Participant Physical and Mental Health and Association with Pandemic Health Impact (n=1775)

|  | n (%) /<br>mean ± SD | Perceived Impact of Pandemic on Health Condition |  |  |  |
| --- | --- | --- | --- | --- | --- |
|  |  | No Effect<br>(n=993) | Minor or<br>Greater<br>(n=782) | X<sup>2</sup>/t | p |
| Chronic Condition |  |  |  |  |  |
| Coronary artery disease | 333 (18.8) | 132 (13.3) | 201 (25.7) | 44.209 | <0.001 |
| Heart failure | 49 (2.8) | 12 (1.2) | 37 (4.7) | 20.228 | <0.001 |
| Arrhythmia | 189 (10.6) | 75 (7.6) | 114 (14.6) | 22.693 | <0.001 |
| Hypertension | 938 (52.8) | 481 (48.4) | 457 (58.4) | 17.559 | <0.001 |
| Cerebrovascular disease | 98 (5.5) | 37 (3.7) | 61 (7.8) | 13.923 | <0.001 |
| Diabetes | 405 (22.8) | 210 (21.1) | 195 (24.9) | 3.565 | 0.059 |
| Renal disease | 62 (3.5) | 23 (2.3) | 39 (5.0) | 9.259 | 0.002 |
| Other | 780 (43.9) | 420 (43.7) | 360 (44.2) | 0.049 | 0.825 |
| Years living with chronic condition |  |  |  |  |  |
| <5 years | 692 (39.0) | 415 (41.8) | 277 (35.4) | 10.488 | 0.015 |
| 5-10 years | 510 (28.7) | 273 (27.5) | 237 (30.3) |  |  |
| 10-20 years | 344 (19.4) | 173 (17.4) | 171 (21.9) |  |  |
| >20 years | 229 (12.9) | 132 (13.3) | 97 (12.4) |  |  |
| History psychiatric disorder |  |  |  |  |  |
| Yes, being treated with meds | 148 (8.3) | 49 (4.9) | 99 (12.7) | 97.857 | <0.001 |
| Yes, but untreated | 116 (6.5) | 29 (2.9) | 87 (11.1) |  |  |
| Yes, but remitted | 97 (5.5) | 45 (4.5) | 52 (6.6) |  |  |
| No | 1414 (79.7) | 870 (87.6) | 544 (69.6) |  |  |
| Major Life Event (n, % yes) | 232 (13.1) | 103 (10.4) | 129 (16.5) | 14.438 | <0.001 |
| Somatic Self-rating Scale (/80) | 36.07±10.50 | 31.38±7.68 | 42.03±10.58 | -23.666 | <0.001 |
| Somatic Symptoms (/36) | 15.54±4.65 | 13.52±3.47 | 18.10±4.69 | -22.864 | <0.001 |
| Anxiety symptoms (/20) | 8.84±2.95 | 7.68±2.18 | 10.30±3.14 | -19.925 | <0.001 |
| Depressive symptoms (/16) | 7.59±2.61 | 6.56±2.12 | 8.90±2.59 | -20.437 | <0.001 |
| Sleep & cognitive symptoms (/8) | 4.10 ± 1.36 | 3.62 ± 1.17 | 4.72 ± 1.35 | -18.017 | <0.001 |
| SCL Depressive Items (/25) | 9.94±4.29 | 8.35 ± 3.28 | 11.95±4.57 | -18.563 | <0.001 |
| Sadness (/5) | 2.39±1.02 | 2.07 ± 0.88 | 2.80 ± 1.03 | -15.703 | <0.001 |
| Suicidal ideation (/5) | 1.45±0.80 | 1.21 ± 0.54 | 1.75 ± 0.96 | -13.882 | <0.001 |
| Hopelessness (/5) | 1.83±1.02 | 1.50 ± 0.79 | 2.25 ± 1.12 | -15.865 | <0.001 |
| Feel empty inside (/5) | 2.04±1.03 | 1.70 ± 0.85 | 2.47 ± 1.07 | -16.400 | <0.001 |
| Anhedonia (/5) | 2.23±1.09 | 1.87 ± 0.93 | 2.69 ± 1.10 | -16.622 | <0.001 |

SD, standard deviation
Note: some items allowed single responses and others multiple.

Mean psychosocial well-being scores are shown in Table 2. Approximately 80% had no known psychiatric history. The 41.5% of participants with elevated SSS scores indicating psychological distress (Table 3) were significantly more likely to be women, older, unmarried, living alone, with no fixed income, and had lower educational attainment than those with subclinical scores (all p ≤ 0.016). All chronic conditions except hypertension and diabetes were associated with significantly more distress on the SSS (all p ≤0.013), and they were living longer with their condition (p < 0.001). They were significantly more likely to have suffered a major life event (16.7% vs. 10.5%; p < 0.001).

**Table 3:** Participant Pandemic Attitudes and Impact on Chronic Disease Management (N=1775)

| | n (%) / mean ± SD | Association with Somatic Symptoms | | | |
| --- | --- | --- | --- | --- | --- |
|  |  | Elevated (n=736) | Subclinical (n=1039) | X<sup>2</sup>/t | P |
| Source of Pandemic-related Information |  |  |  |  |  |
| Traditional and new media (e.g., WeChat) | 1624 (91.5) | 651 (88.5) | 973 (93.6) | 16.124 | < 0.001 |
| Television and newspaper only (traditional media) | 110 (6.2) | 59 (8.0) | 51 (4.9) |  |  |
| No media, only family or neighbors | 41 (2.3) | 36 (3.5) | 15 (1.4) |  |  |
| Impact of Pandemic on Life |  |  |  |  |  |
| None | 210 (11.8) | 42 (5.7) | 168 (16.2) | 200.903 | < 0.001 |
| Only physical distancing | 184 (10.4) | 108 (14.7) | 76 (7.3) |  |  |
| Temporary | 947 (53.4) | 302 (41.0) | 645 (62.1) |  |  |
| Permanent changes | 434 (24.5) | 284 (38.6) | 150 (14.4) |  |  |
| View of Pandemic Status in Future and Control Measures |  |  |  |  |  |
| Situation is serious, and worry about future | 652 (36.7) | 378 (51.4) | 274 (26.4) | 144.777 | < 0.001 |
| Think the strict measures will turn things around | 403 (22.7) | 144 (19.6) | 259 (24.9) |  |  |
| Always optimistic | 397 (22.4) | 85 (11.5) | 312 (30.0) |  |  |
| I am not worried about getting Omicron | 323 (18.2) | 129 (17.5) | 194 (18.7) |  |  |
| How Long Strict Control Measures can be Tolerated |  |  |  |  |  |
| I cannot even endure 1-2 weeks | 166 (9.4) | 111 (15.1) | 55 (5.3) | 65.327 | < 0.001 |
| A month at maximum | 847 (47.7) | 367 (49.9) | 480 (46.2) |  |  |
| 3 months maximum | 258 (14.5) | 79 (10.7) | 179 (17.2) |  |  |
| 6 months maximum | 23 (1.3) | 9 (1.2) | 14 (1.3) |  |  |
| As long as I can get necessities, I can endure it for a long time | 481 (27.1) | 170 (23.1) | 311 (29.9) |  |  |
| Impact of Pandemic on Health Condition† |  |  |  |  |  |
| No effect | 993(55.9) | 214 (29.1) | 779 (75.0) | 407.066 | < 0.001 |
| Minor | 564 (31.8) | 338 (45.9) | 226 (21.8) |  |  |
| Serious | 218 (12.3) | 184 (25.0) | 34 (3.3) |  |  |
| Adherence to Control Measures |  |  |  |  |  |
| Stayed home | 772 (43.5) | 315 (42.8) | 457 (44.0) | 10.330 | 0.016 |
| Only went out to get food and medicine | 518 (29.2) | 193 (26.2) | 325 (31.3) |  |  |
| Community sealed, so could not go out | 460 (25.9) | 215 (29.2) | 245 (23.6) |  |  |
| No change | 25 (1.4) | 13 (1.8) | 12 (1.2) |  |  |
| Exercise Maintenance when Strict Control Measures† |  |  |  |  |  |
| Changed to Exercise in home | 761 (42.9) | 240 (32.6) | 521 (50.1) | 58.656 | < 0.001 |
| Stopped exercise because of pandemic | 595 (33.5) | 306 (41.6) | 289 (27.8) |  |  |
| Do not exercise as usual | 366 (20.6) | 169 (23.0) | 197 (19.0) |  |  |
| Exercise in residential area as before | 53 (3.0) | 21 (2.9) | 32 (3.1) |  |  |
| Access to Medication When Strict Control Measures |  |  |  |  |  |
| I was sure to stock up before | 845 (47.6) | 306 (41.6) | 539 (51.9) | 50.597 | < 0.001 |
| Someone else got my medication | 360 (20.3) | 165 (22.4) | 195 (18.8) |  |  |
| Discontinued use | 310 (17.5) | 170 (23.1) | 140 (13.5) |  |  |
| Internet dispensing | 228 (12.8) | 74 (10.1) | 154 (14.8) |  |  |
| Borrowed medicine from another | 32 (1.8) | 21 (2.9) | 11 (1.1) |  |  |
| Available Supply of Chronic Disease Medication |  |  |  |  |  |
| I discontinued | 236 (13.3) | 116 (15.8) | 120 (11.5) | 66.290 | < 0.001 |
| Not enough even for a week | 309 (17.4) | 168 (22.8) | 141 (13.6) |  |  |
| Enough for 1-2 weeks | 469 (26.4) | 213 (28.9) | 256 (24.6) |  |  |
| Enough for 2-4 weeks | 393 (22.1) | 138 (18.8) | 255 (24.5) |  |  |
| More than a month on hand | 368 (20.7) | 101 (13.7) | 267 (25.7) |  |  |
| Greatest Worries about Chronic Condition Related to Omicron |  |  |  |  |  |
| Being unable to access care due to control measures | 1477 (83.2) | 640 (87.0) | 837 (80.6) | 12.625 | < 0.001 |
| Getting infected and deteriorating | 747 (42.1) | 372 (50.5) | 375 (36.1) | 36.913 | < 0.001 |
| The economic impact of the pandemic affects health resourcing | 438 (24.7) | 190 (25.8) | 248 (23.9) | 0.878 | 0.349 |
| Other | 112 (6.3) | 40 (5.4) | 72 (6.9) | 1.629 | 0.202 |
| What Needed during Waves to Manage Chronic Condition |  |  |  |  |  |
| Easier access to medications | 1165 (65.6) | 478 (64.9) | 687 (66.1) | 0.264 | 0.607 |
| Ensuring access to care regardless of control measures | 1147 (64.6) | 512 (69.6) | 635 (61.1) | 13.451 | < 0.001 |
| Guarantee of emergency hospital and outpatient care access | 992 (55.9) | 450 (61.1) | 542 (52.2) | 14.079 | < 0.001 |
| Facilitating access to care even if I cannot use or do not have a mobile phone | 760 (42.8) | 335 (45.5) | 425 (40.9) | 3.742 | 0.053 |
| Greater availability of telehealth (y) | 659 (37.1) | 265 (36.0) | 394 (37.9) | 0.677 | 0.411 |

Note: some items allowed single responses and others multiple.
SD, standard deviation
†variables used for association tests in previous table

### Impact of Pandemic

As shown in Table 3, most participants perceived the pandemic had a temporary impact on their lives, but almost one-quarter perceived changes would be permanent. In terms of the future, over one-third were very worried about it, and one quarter each were optimistic or perceived the control measures would be successful in mitigating COVID-19 impact. Most commonly, participants reported they could withstand the lockdown for a month.

Half perceived the pandemic had at least a minor impact on their health or more (Table 3). As shown in Table 2, those perceiving health impact were significantly more likely to be older, unmarried, with lower educational attainment and income. Participants with cardiovascular, renal, and psychiatric diseases, who had their diseases for more years, reported major life events, and higher somatic and depressive symptoms (including all subscales) were also significantly more likely to perceive impact of the pandemic on their health than their counterparts (Table 3).

Forty percent exercised in their home, but over one-third stopped exercising and one-fifth changed their exercise (Table 3). Participants who had their condition for fewer years (p = 0.009) and who had lower somatic (p < 0.001) were more likely to change their exercise due the pandemic and associated control measures (no association with chronic condition).

With regard to medication, while almost half tried to stock up before the lockdown, almost one-fifth of participants discontinued use (Table 3). Only just over 10% of this older cohort used an internet-based pharmacy. Of those that still had medication, just under half had enough supply to last at least 2 weeks.

Participant’s greatest worries related to Omicron and their chronic condition are also shown in Table 3. Chief among them were fears around inability to access healthcare and medicines, as well as contracting COVID-19 and their health deteriorating. To best manage their chronic condition during the wave, participants most wanted guaranteed access to care and medication, particularly even if they do not have – or are not able to use – a mobile phone.

Pandemic perceptions were highly associated with SSS (Table 3). Participants who were significantly less likely to use new media to get pandemic-related information and more often relied on family or neighbours for information only, who were more likely to view pandemic impacts on their lives as permanent, were less optimistic about the future, reported being able to tolerate control measures for a significantly shorter amount of time, perceived a significantly greater impact of the pandemic on their health, were less likely to go out even to get food or medicine, were more likely to stop exercise, and were more likely to discontinue medicine use or had less supply on hand, had elevated SSS scores, indicating not only somatic but anxious, depressive and sleep-related symptoms. These participants with elevated distress were also more concerned about all but 2 of the worries related to the pandemic and were more likely to want assurances around access to in- and outpatient care regardless of COVID-19 wave status and associated control measures. An adjusted model is shown in Table 4.

**Table 4:** Model Assessing Correlates of SSS

| **Independent variable** | **OR** | **95% CI** | ***p*** |
| --- | --- | --- | --- |
| Age | 0.997 | 0.985 - 1.008 | 0.572 |
| **Sex** |  |  |  |
| Male | 1 |  |  |
| Female | 1.577 | 1.220 - 2.039 | 0.001 |
| **Marital Status** |  |  |  |
| Married | 1 |  |  |
| Widowed | 0.898 | 0.502 - 1.606 | 0.716 |
| Never married | 0.808 | 0.446 - 1.463 | 0.481 |
| Divorced | 1.196 | 0.633 - 2.259 | 0.581 |
| **Living Arrangement** |  |  |  |
| Alone | 1 |  |  |
| With partner only | 0.678 | 0.424 - 1.086 | 0.106 |
| With children and/or other extended family members | 0.677 | 0.438 - 1.045 | 0.078 |
| **Educational Attainment** |  |  |  |
| Junior high school and below | 1 |  |  |
| Junior college or senior high school | 1.021 | 0.693 - 1.503 | 0.918 |
| Bachelor and above | 0.842 | 0.552 - 1.284 | 0.425 |
| **Income Status** |  |  |  |
| No fixed income (e.g., rely on children, state subsidies) | 1 |  |  |
| Has work income | 0.696 | 0.433 - 1.118 | 0.134 |
| Has additional source of income on top of salary (e.g., online) | 0.630 | 0.330 - 1.203 | 0.161 |
| **Diagnosis** |  |  |  |
| Coronary Artery Disease | 2.006 | 1.439 - 2.797 | < 0.001 |
| Arrhythmia | 2.588 | 1.731 - 3.870 | < 0.001 |
| Pandemic Media Source Type |  |  |  |
| Traditional and new media (e.g., WeChat) | 1 |  |  |
| Television and newspaper only (traditional media) | 0.932 | 0.558 - 1.555 | 0.787 |
| No media, only family or neighbors | 1.139 | 0.432 - 3.002 | 0.793 |
| Perceived Impact of Pandemic on Life |  |  |  |
| None | 1 |  |  |
| Only physical distancing | 2.566 | 1.491 - 4.418 | 0.001 |
| Temporary | 1.433 | 0.929 - 2.209 | 0.103 |
| Permanent changes | 3.298 | 2.051 - 5.305 | < 0.001 |
| Perception of Future & Control Measures |  |  |  |
| Situation is serious, and worry about future | 1 |  |  |
| Think the strict measures will turn things around | 0.632 | 0.456 - 0.876 | 0.006 |
| Always optimistic | 0.418 | 0.291 - 0.600 | < 0.001 |
| I am not worried about getting Omicron | 0.636 | 0.453 - 0.894 | 0.009 |
| Duration can Tolerate Control Measures |  |  |  |
| I cannot even endure 1-2 weeks | 1 |  |  |
| A month at maximum | 0.534 | 0.341 - 0.835 | 0.006 |
| 3 months maximum | 0.415 | 0.245 - 0.706 | 0.001 |
| 6 months maximum | 0.623 | 0.199 - 1.951 | 0.416 |
| As long as I can get necessities, I can endure it for a long time | 0.496 | 0.305 - 0.806 | 0.005 |
| Adherence to Control Measures |  |  |  |
| No change | 1 |  |  |
| Only went out to get food and medicine | 0.854 | 0.301 - 2.423 | 0.767 |
| Stayed home | 0.972 | 0.343 - 2.756 | 0.957 |
| Community sealed, so could not go out | 1.017 | 0.355 - 2.913 | 0.975 |
| Perceived Impact of Pandemic on Health Condition |  |  |  |
| No effect | 1 |  |  |
| Minor | 3.900 | 3.024 - 5.030 | < 0.001 |
| Serious | 12.905 | 8.340 - 19.968 | < 0.001 |
| Change to Exercise Routine due to Pandemic |  |  |  |
| Do not exercise as usual | 1 |  |  |
| Stopped exercise because of pandemic | 0.850 | 0.609 - 1.187 | 0.340 |
| Changed to exercise in home | 0.596 | 0.432 - 0.822 | 0.002 |
| Exercise in residential area as before | 0.938 | 0.445 - 1.981 | 0.868 |

SSS: somatic self-rating scale, including physical disorders, depressive, anxious, sleep and cognitive symptoms as well.

## Discussion

This is the first or one of the first full studies to examine the impact of the COVID-19 pandemic and its associated control measures in a large sample of patients with chronic conditions. It was undertaken in the context of some of the most stringent control measures globally in one of the most densely populous cities in the world where risk of transmission is hence greater,^16,17^ and two years into the pandemic when these patients have thus been coping with isolation, economic impacts as well as changed and inconvenient access to care and treatments for a prolonged period. Despite that few were living alone which could mitigate isolation during the lockdown, consistent with findings in the general population,^18,19^ results show elevated distress, but also negative impacts on chronic disease management.

The burden of distress was quite high, even when considering that rates of psychological distress are higher in those with chronic disease^20,21^ and rates in the Chinese population. Generally, rates of distress decline with age, but the opposite finding was found herein, likely due to the increased risk of severe COVID-19 with age.^22-24^ Many patients were very concerned about the impact of the pandemic on their health and their access to care if needed. Research has shown that much preventive and non-preventive care was avoided due to fear of exposure to SARS-CoV-2, and thus periodic health examinations regarding chronic disease risk factors were likely also missed, likely leaving weight,^25^ blood pressure, lipids, and blood glucose less-optimally managed.

In terms of health management, despite the proven benefits of secondary prevention behaviours such as exercise and medication adherence,^26^ many chronic disease patients discontinued them. Despite the availability of online pharmacies and patient’s low supply of medication, these were not widely used, likely due to low digital literacy or technology access in the elderly.^27,28^ One-third stopped exercising, and another quarter changed their exercise rather than moving their exercise routine home. How outpatients were able to maintain a healthy diet with dwindling food supplies would be another important area for study,^29^ as would be impacts of pandemic-related lockdowns on tobacco access and use in chronic disease patients. Moreover, given the importance of relationship quality to health,^30^ the impact of prolonged lockdown on relationships in chronic disease patients warrants study.

In our earlier examination of the impact of the pandemic and associated control measures on older adults (many of whom similarly had hypertension, CVD or diabetes) in the Spring of 2020,^31^ mean scores on the SSS were somewhat lower at 29/80 vs 36 herein (all mean subscale scores were somewhat lower as well). In terms of pandemic impact on life, only 6% perceived this would be permanent in 2020 vs 25% in 2022. Also in 2020, most participants (47%) reported they could endure strict prevention and control measures for a long time (well over 6 months with another third saying they could endure 6 months); in 2022 however, almost 10% said they could not even endure strict prevention and control measures for a week or two and most (48%) reported they could endure them for a maximum of a month. Also in 2020, 94% reported no effect of the pandemic on their health condition, yet this was only 56% in 2022. Overall, for the common measures across the two surveys, perceptions worsened with time.

Study implications relate to ensuring patients have what they need to manage their condition. What they most wanted was easier access to healthcare and medications, regardless of control measures. This is despite quite fast and broad availability of telehealth in China and online pharmacies.^32^ This could likely be due to lower digital literacy or device availability in older adults, or preferences for in-person services.^33,34^ Efforts to close the age-related “digital divide” must continue. Greater access to virtual cardiac rehabilitation in China could address these issues as well.^6,35^

Caution is warranted when interpreting these results. First, this is a cross-sectional study, so the design precludes causal determinations. Second, the results may not be generalizable beyond China with its political and cultural context, and the city of Shanghai more specifically where the stringency of control measures was high. Nevertheless, the population of Shanghai is very large, and findings can inform other governments when considering their control measures. Moreover, similarity of the sample to the larger population is not known given the recruitment strategy, designed to quickly secure data during the lockdown, but where full reach and number of non-respondents were not collated. Third, all data were self-report, and hence there may have been socially desirable responding or other measurement error, and psychiatric conditions nor health status were not verified through structured clinical interview or medical records. Fourth, due to the exploratory nature of the study, multiple comparisons were performed, increasing the chance of type 1 error. Finally, whether patients had COVID-19 prior to or during the period of study was not considered; more research is needed.

In conclusion, two years into the pandemic in the midst of arguably the most stringent prevention and control measures globally, over 40% of chronic disease patients are experiencing elevated psychosocial distress, and this was associated with female, diagnosis of coronary artery disease and arrhythmia, perceived impact of pandemic on life, and impact of pandemic on health condition (Figure 1). Many patients stopped exercise and discontinued medication, and their greatest fears concerned access to healthcare. They were less able to tolerate prevention and control measures and had more negative perceptions related to the pandemic than at its beginning. Greater access to cardiac rehabilitation in China, including delivery supported by technology where patients are able, could address these issues.

**Figure 1:**
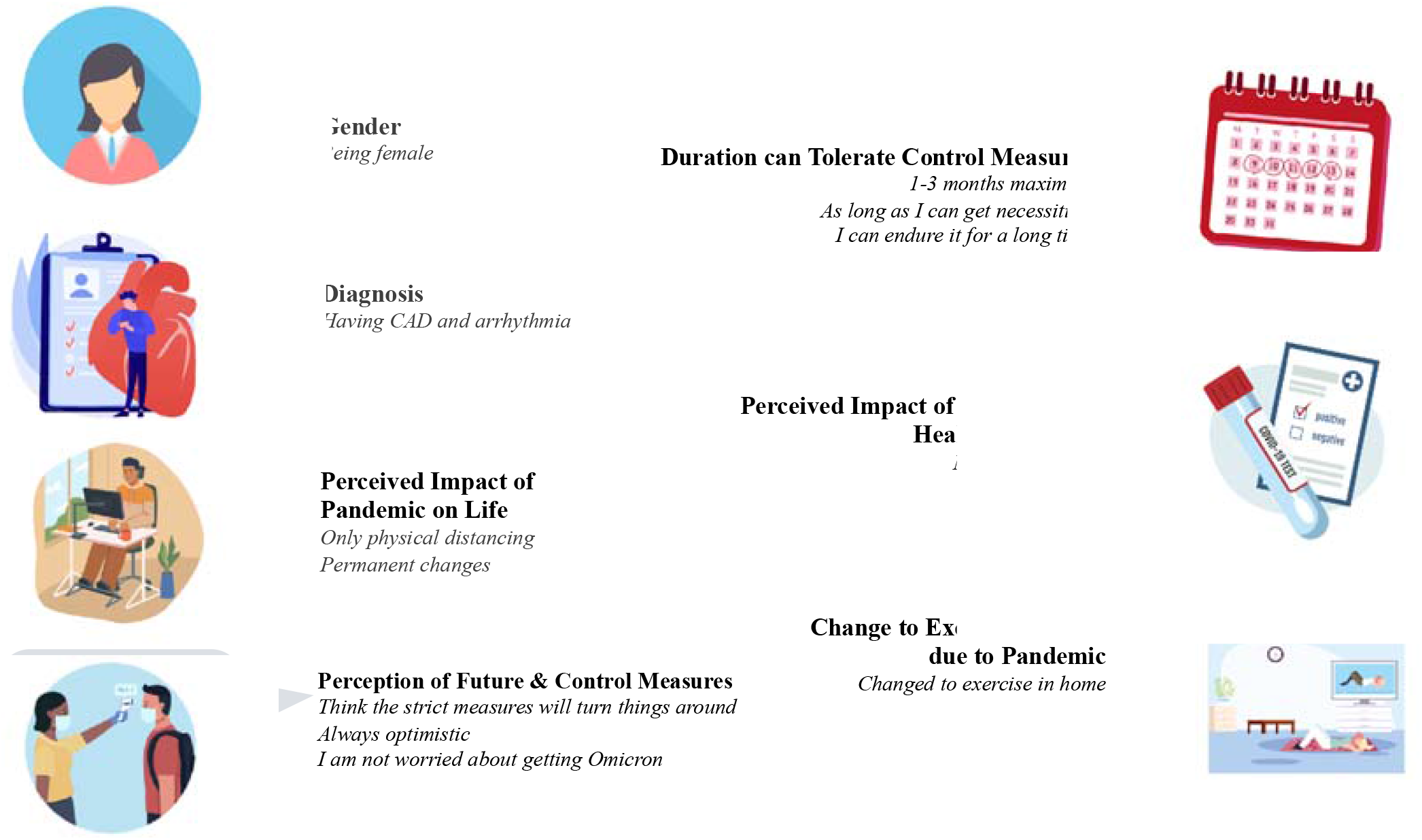
Correlates of Psychosocial Distress in Chronic Disease Patients during Stringent COVID-19 Prevention and Control Measures CAD: coronary artery disease

## Data Availability

All data produced in the present study are available upon reasonable request to the authors

